# Comprehensive validation of fasting- and oral glucose tolerance test-based indices of insulin secretion against gold-standard measures

**DOI:** 10.1101/2022.04.14.22273491

**Authors:** Katsiaryna Prystupa, Rebecka Renklint, Youssef Chninou, Julia Otten, Louise Fritsche, Sebastian Hörber, Andreas Peter, Andreas Birkenfeld, Andreas Fritsche, Martin Heni, Robert Wagner

## Abstract

**Background and aims:** With prediabetes and diabetes increasingly recognized as heterogenous conditions, assessment of beta-cell function is gaining clinical importance to identify disease subphenotypes. Our study aims to comprehensively validate all types of surrogate indices based on OGTT- and fasting-measurements in comparison with gold standard methods.

**Materials and methods:** The hyperglycaemic clamp extended with GLP-1 infusion and IVGTT, as well as OGTT, was performed in two well-phenotyped cohorts. The gold-standard-derived indices were compared with surrogate insulin secretion markers, derived from fasting state and OGTT, using both Pearson’s and Spearman’s correlation coefficients. The insulin- and C-peptide-based indices were analysed separately in different groups of glucose tolerance and the entire cohorts.

**Results:** The highest correlation coefficients were found for AUC (I_0-30_)/AUC (G_0-30_), first-phase Stumvoll and Kadowaki model. These indices have high correlation coefficients with measures obtained from both insulin and C-peptide levels from IVGTT and hyperglycaemic clamp. AUC (I_0-30_)/AUC (G_0-30_), first-phase Stumvoll, AUC (I_0-120_)/AUC (G_0-120_) and BIGTT-AIR_0-60-120_ demonstrated the strongest association with incretin-stimulated insulin response.

**Conclusion:** We have identified glucose- and GLP-1-stimulated insulin secretion indices, derived from OGTT and fasting state, that have the strongest correlation with gold-standard measures and could be potentially used in future researches and clinical practice.

## INTRODUCTION

Reduced sensitivity to the action of insulin and deficient insulin secretion are two physiological defects underlying type 2 diabetes and impaired glucose tolerance. Insulin secretion and sensitivity are associated across a negative feedback loop, in which beta-cells reimburse for changes in whole-body insulin sensitivity through a proportional and reciprocal increase in insulin secretion^1^.

Insulin sensitivity and insulin secretion can be accurately determined by hyperinsulinemic-euglycemic and hyperglycaemic clamps or intravenous glucose tolerance tests (IVGTT) that are considered “gold standards” for these measurements ^2,3^. Precise assessment of these two traits is crucial to identify subphenotypes of prediabetes and type 2 diabetes, which is gaining increasing importance by acknowledgment of the pathophysiologic heterogeneity of this condition^4,5^. Furthermore, these measures can support the prediction of diabetes in non-diabetic subjects^6,7^. However, the invasive, expensive and time-consuming gold-standard procedures are not applicable in routine clinical practice.

Several indices have been proposed to estimate insulin sensitivity and insulin secretion, based on more readily measurable parameters obtained in the fasting state or after an oral glucose tolerance test (OGTT)^8^. Their usefulness is influenced by the degree of correlation with gold standard indicators, by their reproducibility^9^, and by their ability to predict diabetes incidence similarly to more complex methods ^10,11,12^.

Many surrogate methods for insulin sensitivity assessment have been proposed and validated^10,12^, but comprehensive studies to compare insulin secretion measures to gold standard methods are scarce. While several indices for beta-cell assessment based on dynamic changes in insulin and glucose during OGTT have been separately evaluated in numerous studies, most of these indices have not been compared head-to-head and there is still disagreement about their validity^13,14,15^. Therefore, our work is aimed at performing a large, comprehensive comparative analysis of all types of indicators obtained in the fasting state or during OGTT to determine the secretion of insulin in comparison with the gold standards IVGTT and hyperglycemia clamp in two well-phenotyped cohorts. Moreover, the novel hyperglycaemic clamp was used in one of our included trials^16^ allows for defining surrogate indices for GLP1-stimulated insulin secretion.

## RESEARCH DESIGN AND METHODS

### Participants

The 392 subjects included in the present analysis were part of the Tuebingen Family Study (TUEF), who were recruited based on their increased risk for type 2 diabetes (prior gestational diabetes, overweight or positive family history). 316 subjects participated in OGTT and IVGTT, 76 in OGTT and hyperglycaemic clamps. Informed written consent to the studies was obtained from all participants. The studies adhered to the Declaration of Helsinki. The study protocols were approved by the Ethical Committee of the Medical Faculty of the University of Tübingen.

All participants were classified into different stages of glucose tolerance according to the revised World Health Organization criteria^26^. The IVGTT-cohort consists of 209 subjects with normal glucose tolerance (NGT), 91 with impaired fasting glucose (IFG) and/or impaired glucose tolerance (IGT), and 16 with test-diagnosed type 2 diabetes (Table 1). Of the 91 subjects with IFG/IGT, 28 had isolated IFG (fasting plasma glucose 6.1–6.9 mmol/l), 40 - isolated IGT (2-h glucose value 7.8–11.0 mmol/l), 23 -both IFG and IGT, 16 - newly diagnosed type 2 diabetes (T2D). The Hyperglycaemic clamp group comprises 49 NGT, 23 prediabetes (IFG and/or IGT) and 4 screen-diagnosed T2D participants.

**Table 1.**
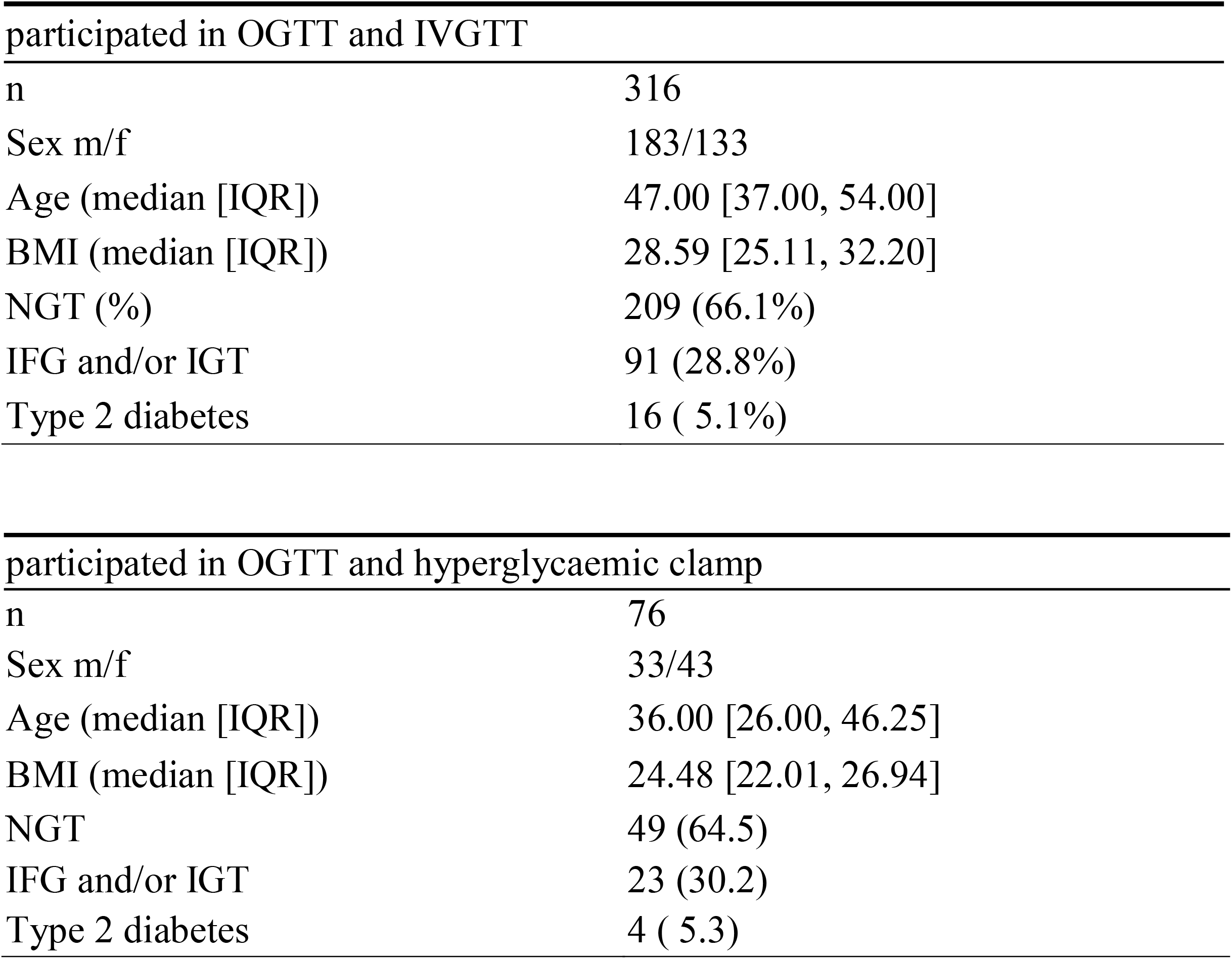
Clinical and biochemical characteristics of the study population.

### Procedures and calculations

In the IVGTT, an intravenous bolus injection of glucose was given (0.3 g/kg body weight in a 20% solution) at time 0 after baseline blood draw. Blood was sampled at 0, 2, 4, 6, 8, 10, 20, 30, 40, 50 and 60 min for measuring plasma glucose, insulin, and C-peptide (CP) concentration^17^. The incremental area under curve (iAUC_0-10_) for insulin and C-peptide over the first 10 min and 10-60 min of the test was determined using the trapezoidal method. Multivariate imputation by chained equations^18^ was performed for missingness, which was less than 2.6% and completely random.

The hyperglycaemic clamp was performed as previously described^16^. In brief, the clamp was initiated with a weight-adapted intravenous bolus of 20% glucose over 1 min, and continued with an infusion of 20% glucose with periodic adjustments based on the negative feedback principle to maintain blood glucose at 10 mmolL^-1^. After 120 min, GLP-1 was given as a bolus injection (4.5 pmol kg^-1^), followed by continuous infusion of 1.5 pmol kg^-1^min^-1^during the next 60 min. Serum samples for insulin, C-peptide, and glucose were drawn at -30, -15, 0, 2.5, 5, 7.5, 10, 15, 30, 60, 90, 120, 140, 160, 170, 180 min. Insulin secretion after an arginine bolus given at the end of the clamp was not analysed in the current work. First phase insulin release, reflecting the early insulin peak secreted from the pancreatic beta-cell in response to glucose stimulation, was calculated as the sum of insulin or C-peptide measurements at 2.5, 5, 7.5 and 10 min. The second phase of glucose-stimulated insulin release was derived as means of insulin or C-peptide at 80, 100 and 120 min. GLP-1-stimulated insulin (C-peptide) secretion at 10 mM glucose was assessed as a mean of 160, 170 and 180 min.

C-peptide and insulin concentrations were determined using chemiluminescent methods on an ADVIA Centaur XPT analyzer (Siemens Healthineers, Eschborn, Germany). Glucose concentrations were measured using a hexokinase method on an ADVIA clinical chemistry XPT analyzer (Siemens Healthineers) All 392 persons participated in the OGTT, taking 75 g of glucose in a volume of 300 ml after an overnight fast. Samples for glucose and insulin measurements were taken at 0, 30, 60, and 120 minutes. Insulin secretion indices were calculated from these OGTTs.

The insulinogenic index (IGI), an index of beta-cell function, was computed as (I_30_ − I_0_)/(G_30_ − G_0_) and (I_120_ − I_0_)/(G_120_ − G_0_), with I_n_, G_n_, plasma concentrations at the nth minute for insulin and glucose respectively^15^.

The beta-cell function insulin sensitivity glucose tolerance test (BIGTT) method is an estimation of an acute insulin response (AIR) derived from the following equation: BIGTT-AIR_0-30-120_ = exp[8.20 + (0.00178 × I_0_) + (0.00168 × I_30_) - (0.000383 × I_120_) - (0.314 × G_0_)- (0.109 × G_30_) + (0.0781 × G_120_) + (0.180 ×sex) + (0.032 × BMI)], BIGTT-AIR_0-60-120_ = exp[8.20 + (0.00178 × I_0_) + (0.00168 × I_60_) - (0.000383 × I_120_) - (0.314 × G_0_)-(0.109 × G_60_) + (0.0781 × G_120_) + (0.180 ×sex) + (0.032 × BMI)]^19^.

The ratio of the AUC for insulin and for C-peptide to AUC for glucose over a specified time frame was calculated by applying the trapezoid rule (AUC (I_all_), AUC (CP_all_), AUC (I_0- 30_)/AUC (G_0-30_), AUC (CP_0-30_)/AUC (G_0-30_), AUC (I_0-120_)/AUC (G_0-120_), AUC (CP_0-120_)/AUC (G_0-120_))^14^. The additional indices of insulin secretion used in this study were as follows: the corrected insulin response (CIR) = I_30_ /(G_30_ x (G_30_ – 3.89)) and I_120_ /(G_120_ x (G_120_ – 3.89))^13^; the insulin/glucose ratio derived as I_0_/G_0_ and I_120_/G_120,_ C-peptide/ glucose ratio CP_0_/G_0_ and CP_120_/G_120_, insulin and C-peptide ratio at minute 30 (I_30_/I_0_, CP_30_/CP_0_) and at minute 120 (I_120_/I_0,_ CP_120_/CP_0_), delta insulin at 30 min ΔI_30_= I_30_ - I_0,_ C-peptide and insulin at fasting state (CP_0_, I_0_), minute 30 (CP_30_, I_30_) and minute 120 (CP_120_, I_120_)^13,20^; first-phase Stumvoll□=□1283□+□1.829□×□I_30_–138.7□×□G_30_+□3.772□×□I_0_; and second-phase Stumvoll□=□286□+□0.416□×□I_30_–25.94□×□G_30_+□0.926□×□I_0_ ^11^; Kadowaki model = (I_30_ − I_0_)/(G_30_ − G_0_)^21^; C-peptide index (CPI) CPI_120_ = 100 × CP_120_(ng/mL)/G_120_(mg/dL), CPI_0_ = 100 × CP_0_(ng/mL)/G_0_(mg/dL)^22^; log transformed insulin at minute 0,30,60,90,120 (log(I_0_), log(I_30_), log(I_60_), log(I_90_), log(I_120_))^23^.

The HOMA-%B was calculated using the fasting plasma insulin and glucose concentration (HOMA-%B = (20 × I_0_)/(G_0_− 3.5))^24^.

Fasting C-peptide, insulin and glucose levels were used in the homeostasis model assessment computer model to generate estimates of beta-cell function (HOMA2%B)^24^.

### Statistical analysis

Statistical analyses were performed with R (software version 4.0.3)^25^. Descriptive data are expressed as medians ± IQR. The relationship between the different estimates of insulin secretion was determined using both Pearson’s and Spearman’s correlation coefficients. The insulin and C-peptide-derived insulin secretion indices were separately analysed. The best-performing indices were selected according to Pearson’s correlation coefficients. P values < 0.05 were considered statistically significant.

## RESULTS

Our analysis included all participants of the TUEF study from whom hyperglycaemic clamp or IVGTT and OGTT were available (N=392). We analyzed data from 316 persons (133 males and 183 females), who underwent intravenous glucose tolerance tests and oral glucose tolerance tests, as well as 76 (33 males and 43 females) in whom hyperglycaemic clamp and OGTT were performed. The characteristics of the study population are summarized in Table 1.

### Comparison of insulin secretion indices with dynamic insulin secretion measurements by IVGTT

The insulin secretion indices obtained from fasting or OGTT measurements were tested against IVGTT-obtained measurements (Insulin iAUC_0-10_ and C-peptide iAUC_0-10_) of beta-cell capacity.

The vast majority of tested surrogate insulin secretion indices except for CP_120_/CP_0_ and I_120_/I_0_ correlated significantly with first-phase insulin and C-peptide-based IVGTT-values in the group of NGT as well as IFG and/or IGT -group (Table 2, Table 3). In NGT, the strongest correlation with 1st-phase insulin and C-peptide by both Spearman’s and Pearson’s methods was found for CIR_30_, BIGTT-AIR_0-30-120_, first-phase Stumvoll and AUC (I_0-30_)/AUC (G_0-30_) (Figure 1, Table 2).

**Table 2:** NGT-group(n=209). Correlation between fasting and OGTT-derived insulin secretion indices and first-phase insulin secretion measured with the IVGTT. (* p<0.05; **p<0.01; ***p<0.001)

| **Indices** | **Insulin**<br>**iAUC<sub>0-10</sub>**<br>**Spearman** | **C-peptide**<br>**iAUC<sub>0-10</sub>**<br>**Spearman** | **Insulin**<br>**iAUC<sub>0-10</sub>**<br>**Pearson** | **C-peptide**<br>**iAUC<sub>0-10</sub>**<br>**Pearson** |
| --- | --- | --- | --- | --- |
| AUC (CP<sub>0-120</sub>)/AUC (G<sub>0-120</sub>) | 0.428 *** | 0.475 *** | 0.392 *** | 0.445 *** |
| AUC (CP<sub>0-30</sub>)/AUC (G<sub>0-30</sub>) | 0.546 *** | 0.572 *** | 0.556 *** | 0.58 *** |
| AUC (CP<sub>all</sub>) | 0.311 *** | 0.296 *** | 0.297 *** | 0.3 *** |
| AUC (I<sub>all</sub>) | 0.435 *** | 0.355 *** | 0.439 *** | 0.373 *** |
| AUC (I<sub>0-120</sub>)/AUC (G<sub>0-120</sub>) | 0.581 *** | 0.522 *** | 0.602 *** | 0.538 *** |
| AUC (I<sub>0-30</sub>)/AUC (G<sub>0-30</sub>) | 0.635 *** | 0.576 *** | 0.665 *** | 0.604 *** |
| BIGTT-AIR<sub>0-30-120</sub> | 0.651 *** | 0.596 *** | 0.697 *** | 0.619 *** |
| BIGTT-AIR<sub>0-60-120</sub> | 0.531 *** | 0.5 *** | 0.54 *** | 0.534 *** |
| CIR<sub>120</sub> | 0.259 *** | 0.255 *** | 0.129 | 0.155 * |
| CIR<sub>30</sub> | 0.646 *** | 0.642 *** | 0.616 *** | 0.616 *** |
| CP<sub>120</sub>/CP<sub>0</sub> | -0.109 | -0.105 | -0.13 | -0.118 |
| CP<sub>30</sub>/CP<sub>0</sub> | 0.264 *** | 0.301 *** | 0.201 ** | 0.241 *** |
| CP<sub>0</sub> | 0.31 *** | 0.278 *** | 0.342 *** | 0.323 *** |
| CP<sub>0</sub>/G<sub>0</sub> | 0.314 *** | 0.294 *** | 0.355 *** | 0.344 *** |
| CP<sub>120</sub> | 0.183 ** | 0.164 * | 0.156 * | 0.152 * |
| CP<sub>120</sub>/G<sub>120</sub> | 0.203 ** | 0.225 ** | 0.179 ** | 0.21 ** |
| CP<sub>30</sub> | 0.52 *** | 0.523 *** | 0.484 *** | 0.496 *** |
| HOMA-%B | 0.439 *** | 0.357 *** | 0.483 *** | 0.417 *** |
| HOMA2-%B (CP) | 0.286 *** | 0.284 *** | 0.313 *** | 0.32 *** |
| HOMA2-%B (I) | 0.401 *** | 0.333 *** | 0.475 *** | 0.404 *** |
| I<sub>0</sub>/G<sub>0</sub> | 0.468 *** | 0.375 *** | 0.517 *** | 0.425 *** |
| I<sub>120</sub> | 0.322 *** | 0.254 *** | 0.297 *** | 0.228 *** |
| I<sub>120</sub>/G<sub>120</sub> | 0.352 *** | 0.296 *** | 0.339 *** | 0.28 *** |
| I<sub>30</sub> | 0.586 *** | 0.519 *** | 0.582 *** | 0.524 *** |
| ΔI<sub>30</sub> | 0.579 *** | 0.517 *** | 0.567 *** | 0.515 *** |
| IGI<sub>120</sub> | 0.222 ** | 0.164 * | 0.136 | 0.109 |
| IGI<sub>30</sub> | 0.665 *** | 0.655 *** | 0.583 *** | 0.594 *** |
| I<sub>120</sub>/I<sub>0</sub> | -0.071 | -0.065 | -0.033 | -0.022 |
| I<sub>30</sub>/I<sub>0</sub> | 0.255 *** | 0.272 *** | 0.193 ** | 0.242 *** |
| Kadowaki model | 0.635 *** | 0.588 *** | 0.662 *** | 0.613 *** |
| log(I<sub>0</sub>) | 0.456 *** | 0.356 *** | 0.491 *** | 0.398 *** |
| log(I<sub>120</sub>) | 0.322 *** | 0.254 *** | 0.325 *** | 0.257 *** |
| log(I<sub>30</sub>) | 0.586 *** | 0.519 *** | 0.572 *** | 0.523 *** |
| log(I<sub>60</sub>) | 0.326 *** | 0.255 *** | 0.305 *** | 0.267 *** |
| log(I<sub>90</sub>) | 0.204 ** | 0.133 | 0.209 ** | 0.132 |
| CPI<sub>0</sub> | 0.314 *** | 0.294 *** | 0.355 *** | 0.344 *** |
| CPI<sub>120</sub> | 0.203 ** | 0.225 ** | 0.179 ** | 0.21 ** |
| first-phase Stumvoll | 0.658 *** | 0.615 *** | 0.686 *** | 0.631 *** |
| second-phase Stumvoll | 0.561 *** | 0.479 *** | 0.574 *** | 0.504 *** |

**Table 3:** IFG and/or IGT -group (n=91). Correlation between fasting and OGTT-derived insulin secretion indices and first-phase insulin secretion measured with the IVGTT. (* p<0.05; **p<0.01; ***p<0.001)

| Indices | Insulin<br>iAUC<sub>0-10</sub><br>Spearman | C-peptide<br>iAUC<sub>0-10</sub><br>Spearman | Insulin<br>iAUC<sub>0-10</sub><br>Pearson | C-peptide<br>iAUC<sub>0-10</sub><br>Pearson |
| --- | --- | --- | --- | --- |
| AUC (CP<sub>0-120</sub>)/AUC (G<sub>0-120</sub>) | 0.573 *** | 0.551 *** | 0.501 *** | 0.513 *** |
| AUC (CP<sub>0-30</sub>)/AUC (G<sub>0-30</sub>) | 0.597 *** | 0.56 *** | 0.555 *** | 0.544 *** |
| AUC (CP<sub>all</sub>) | 0.453 *** | 0.42 *** | 0.426 *** | 0.407 *** |
| AUC (I<sub>all</sub>) | 0.48 *** | 0.367 *** | 0.355 *** | 0.252 * |
| AUC (I<sub>0-120</sub>)/AUC (G<sub>0-120</sub>) | 0.519 *** | 0.435 *** | 0.388 *** | 0.304 ** |
| AUC (I<sub>0-30</sub>)/AUC (G<sub>0-30</sub>) | 0.634 *** | 0.537 *** | 0.513 *** | 0.424 *** |
| BIGTT-AIR<sub>0-30-120</sub> | 0.483 *** | 0.365 *** | 0.411 *** | 0.295 ** |
| BIGTT-AIR<sub>0-60-120</sub> | 0.468 *** | 0.347 *** | 0.51 *** | 0.424 *** |
| CIR<sub>120</sub> | 0.459 *** | 0.433 *** | 0.17 | 0.198 |
| CIR<sub>30</sub> | 0.565 *** | 0.496 *** | 0.452 *** | 0.416 *** |
| CP<sub>120</sub>/CP<sub>0</sub> | -0.128 | -0.079 | -0.169 | -0.11 |
| CP<sub>30</sub>/CP<sub>0</sub> | 0.321 ** | 0.366 *** | 0.274 ** | 0.303 ** |
| CP<sub>0</sub> | 0.331 ** | 0.272 ** | 0.345 *** | 0.294 ** |
| CP<sub>0</sub>/G<sub>0</sub> | 0.356 *** | 0.293 ** | 0.367 *** | 0.319 ** |
| CP<sub>120</sub> | 0.277 ** | 0.249 * | 0.213 * | 0.206 |
| CP<sub>120</sub>/G<sub>120</sub> | 0.452 *** | 0.423 *** | 0.312 ** | 0.326 ** |
| CP<sub>30</sub> | 0.614 *** | 0.575 *** | 0.578 *** | 0.56 *** |
| HOMA-%B | 0.452 *** | 0.309 ** | 0.421 *** | 0.308 ** |
| HOMA2-%B (CP) | 0.382 *** | 0.323 ** | 0.369 *** | 0.333 ** |
| HOMA2-%B (I) | 0.421 *** | 0.282 ** | 0.419 *** | 0.308 ** |
| I<sub>0</sub>/G<sub>0</sub> | 0.438 *** | 0.308 ** | 0.446 *** | 0.343 *** |
| I<sub>120</sub> | 0.289 ** | 0.206 | 0.232 * | 0.134 |
| I<sub>120</sub>/G<sub>120</sub> | 0.385 *** | 0.308 ** | 0.272 ** | 0.19 |
| I<sub>30</sub> | 0.63 *** | 0.535 *** | 0.49 *** | 0.401 *** |
| ΔI<sub>30</sub> | 0.63 *** | 0.539 *** | 0.478 *** | 0.397 *** |
| IGI<sub>120</sub> | 0.43 *** | 0.363 ** | 0.205 | 0.162 |
| IGI<sub>30</sub> | 0.606 *** | 0.528 *** | 0.495 *** | 0.445 *** |
| I<sub>120</sub>/I<sub>0</sub> | -0.068 | -0.057 | -0.097 | -0.075 |
| I<sub>30</sub>/I<sub>0</sub> | 0.248 * | 0.284 ** | 0.164 | 0.188 |
| Kadowaki model | 0.629 *** | 0.548 *** | 0.501 *** | 0.433 *** |
| log(I<sub>0</sub>) | 0.431 *** | 0.298 ** | 0.436 *** | 0.333 ** |
| log(I<sub>120</sub>) | 0.287 ** | 0.206 | 0.207 * | 0.153 |
| log(I<sub>30</sub>) | 0.63 *** | 0.535 *** | 0.56 *** | 0.493 *** |
| log(I<sub>60</sub>) | 0.479 *** | 0.371 *** | 0.435 *** | 0.368 *** |
| log(I<sub>90</sub>) | 0.307 ** | 0.184 | 0.238 * | 0.153 |
| CPI<sub>0</sub> | 0.356 *** | 0.293 ** | 0.367 *** | 0.319 ** |
| CPI<sub>120</sub> | 0.452 *** | 0.423 *** | 0.312 ** | 0.326 ** |
| first-phase Stumvoll | 0.591 *** | 0.498 *** | 0.495 *** | 0.406 *** |
| second-phase Stumvoll | 0.608 *** | 0.497 *** | 0.494 *** | 0.392 *** |

**Figure 1.**
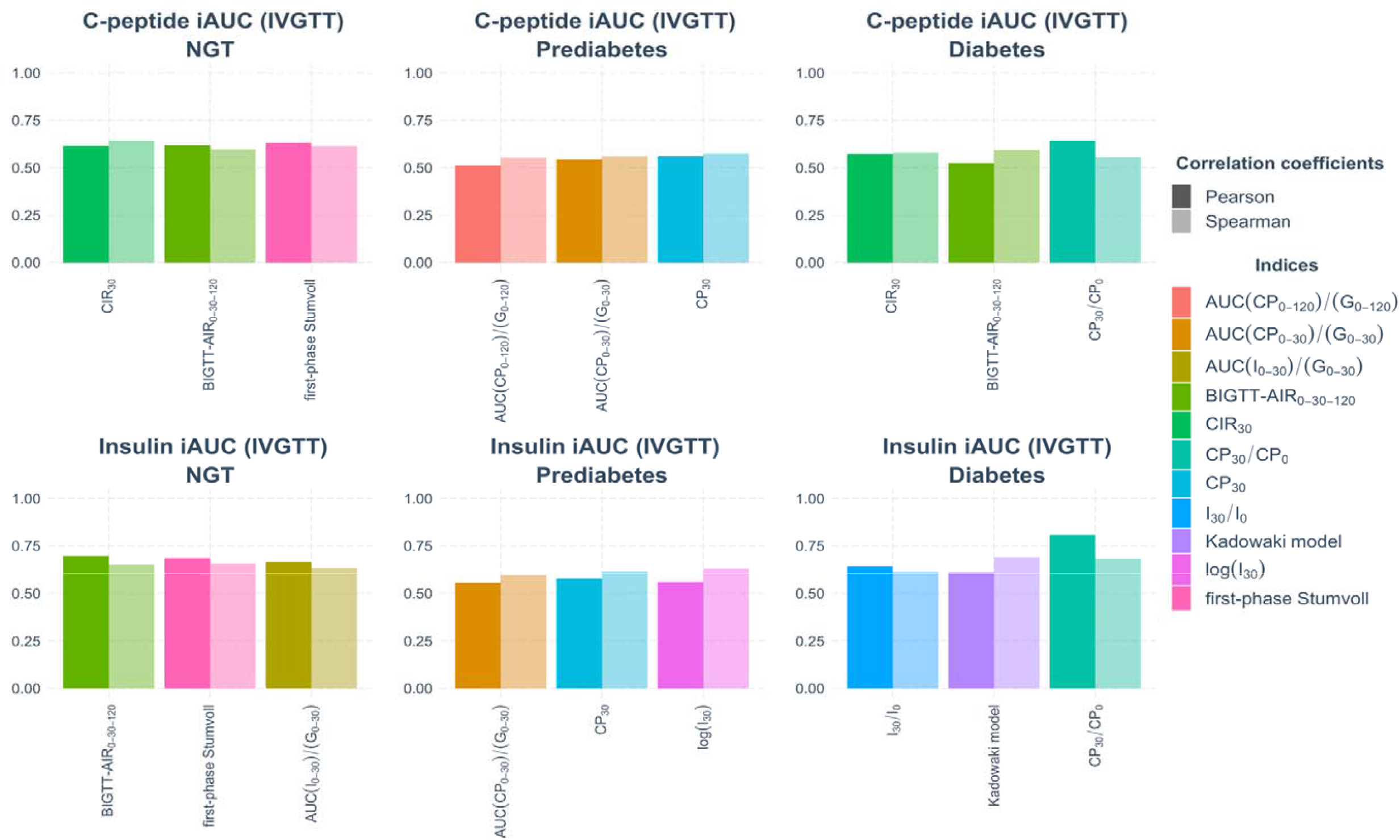
Top-3 indices by Spearman’s and Pearson’s correlation coefficients in subsets with T2D, prediabetes and NGT compared to first-phase IVGTT

In prediabetes, the strongest correlation between Insulin iAUC_0-10_ and C-peptide iAUC_0-10_ was identified for AUC (CP_0-30_)/AUC (G_0-30_), CP_30_, log(I_30_) and AUC (CP_0-120_)/AUC (G_0-120_) (Figure 1, Table 3).

The highest correlation in the entire cohort was detected for AUC (I_0-30_)/AUC (G_0-30_), first-phase Stumvoll, Kadowaki model, IGI_30_ and CIR_30_ (Figure 2). The weakest correlation for both groups had CIR_120_ and IGI_120_.

**Figure 2.**
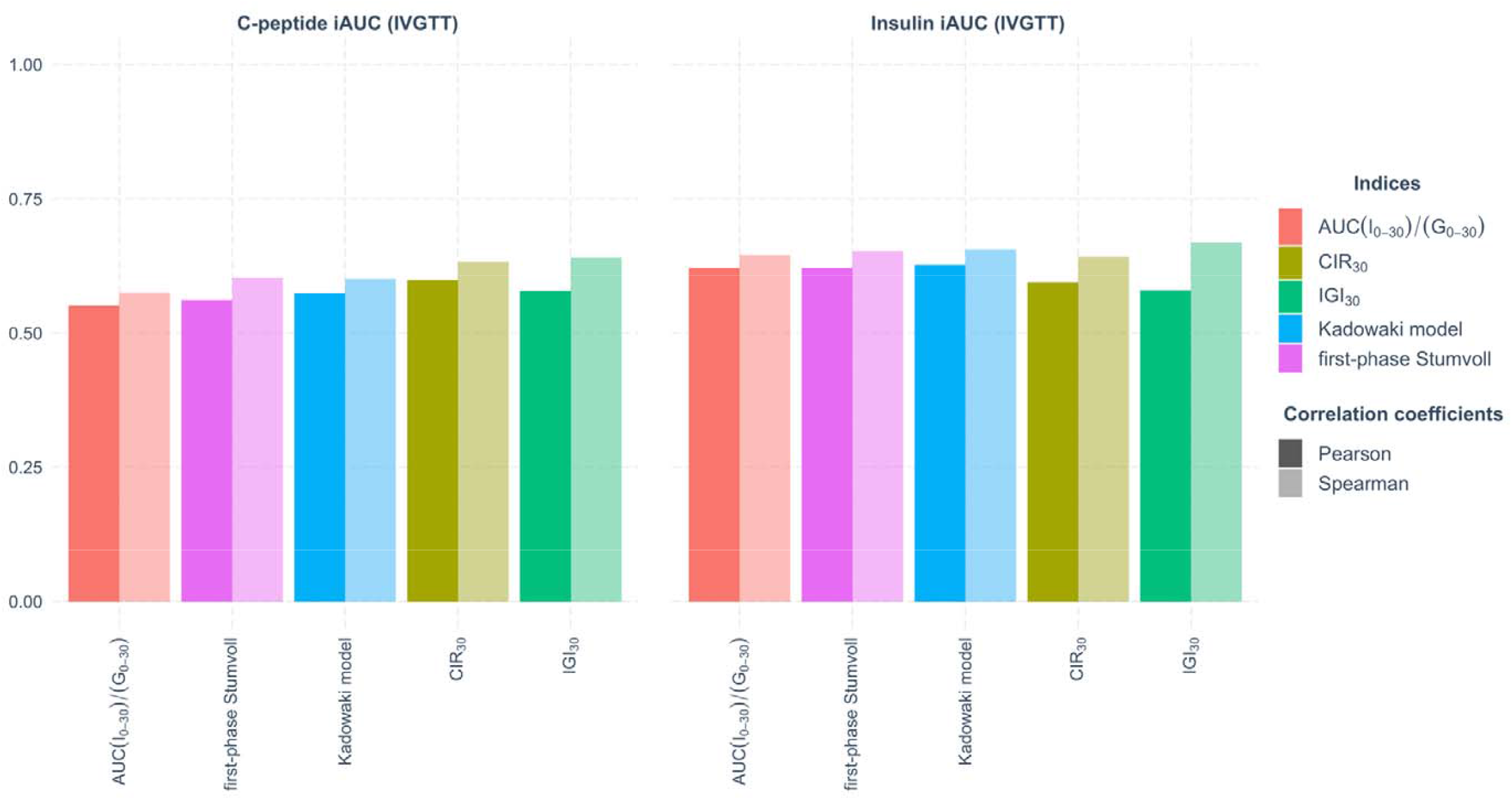
Top indices by Spearman’s and Pearson’s correlation coefficients in the entire cohort (n=316) compared to first-phase IVGTT

We also analysed the correlation coefficients in a group of participants with test-diagnosed type 2 diabetes (n=16). According to our analysis, CP_30_/CP_0_, CIR_30_, I_30_/I_0_, BIGTT-AIR_0-30-120_ and Kadowaki correlated significantly with both Insulin iAUC_0-10_ and C-peptide iAUC_0-10_ (Figure 1, Table 4).

**Table 4:**
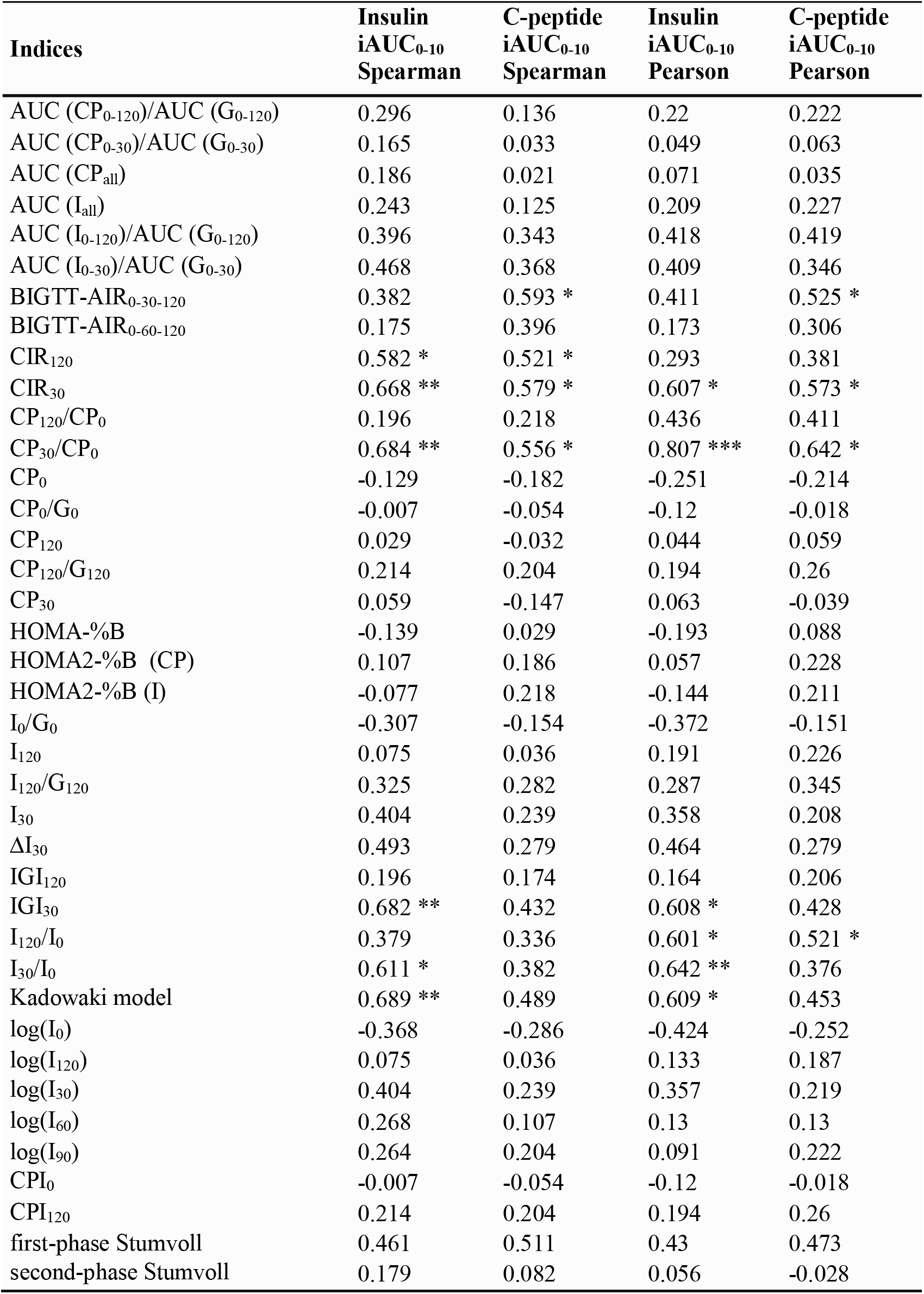
Diabetes -group (n=16). Correlation between fasting and OGTT-derived insulin secretion indices and first-phase insulin secretion measured with the IVGTT. (* p<0.05; **p<0.01; ***p<0.001)

### Comparison of insulin secretion indices with dynamic insulin secretion measurements in the hyperglycaemic clamp

We tested different formulas for calculating insulin secretion from OGTT and hyperglycaemic clamp techniques.

An overwhelming majority of surrogate indices have a positive correlation with the first-phase insulin response obtained from the gold standard in participants with normal glucose tolerance (Table 5.1). In NGT, the numerically highest significant correlation with clamp-derived first-phase insulin secretion measured by insulin and C-peptide was detected for AUC (I_0-120_)/AUC (G_0-120_), AUC (I_0-30_)/AUC (G_0-30_), I_30_ and BIGTT-AIR_0-30-120_ (Figure 3, Table 5.1).

**Table 5.1:**
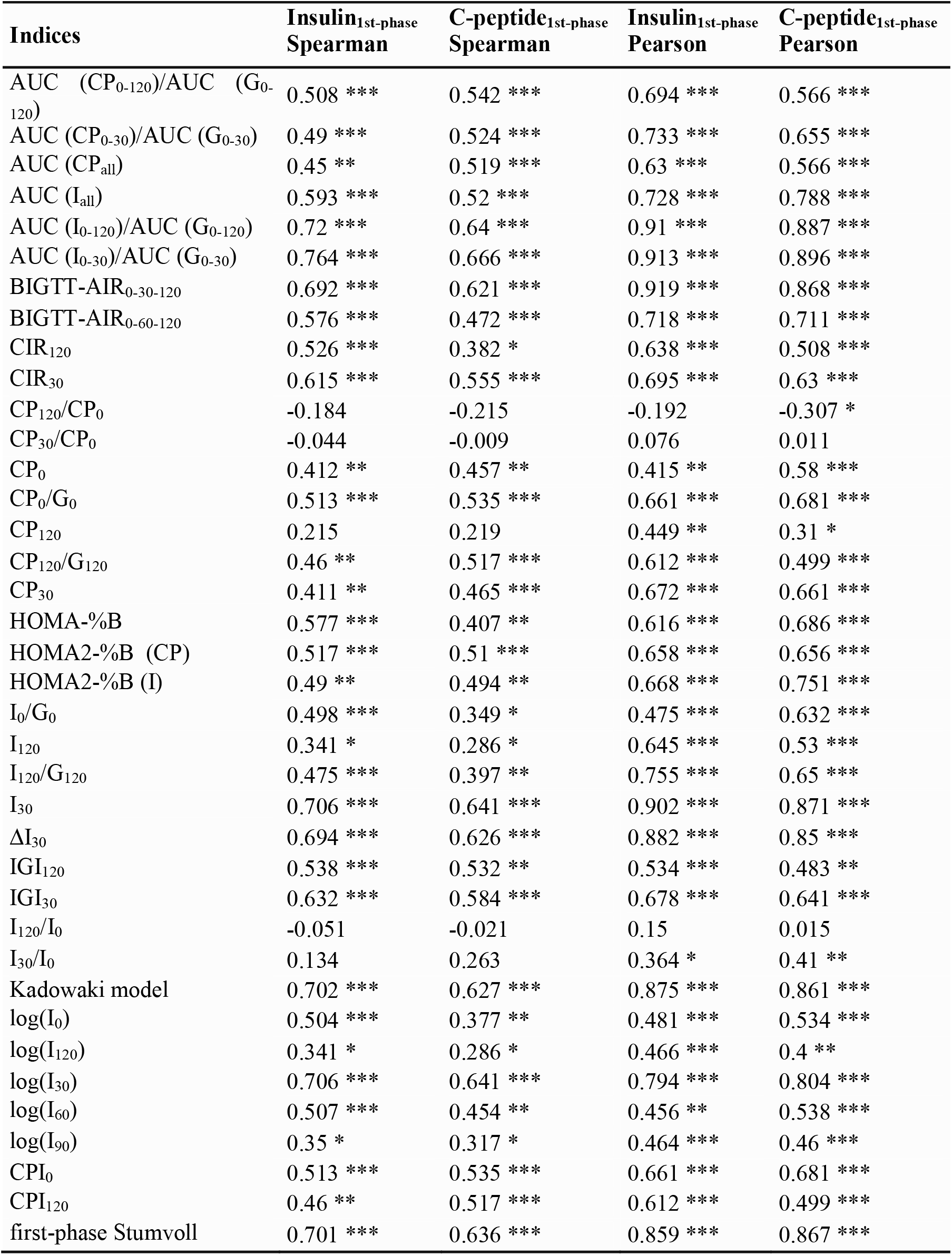
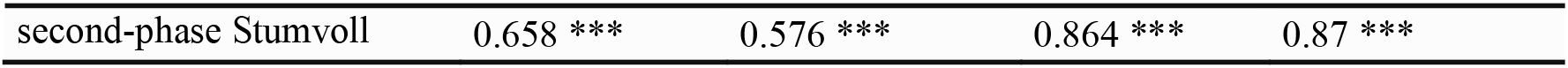
NGT-group(n=49). Correlation between fasting and OGTT-derived insulin secretion indices and first-phase insulin secretion measured with the hyperglycaemic clamp. (* p<0.05; **p<0.01; ***p<0.001)

**Table 5.2.**
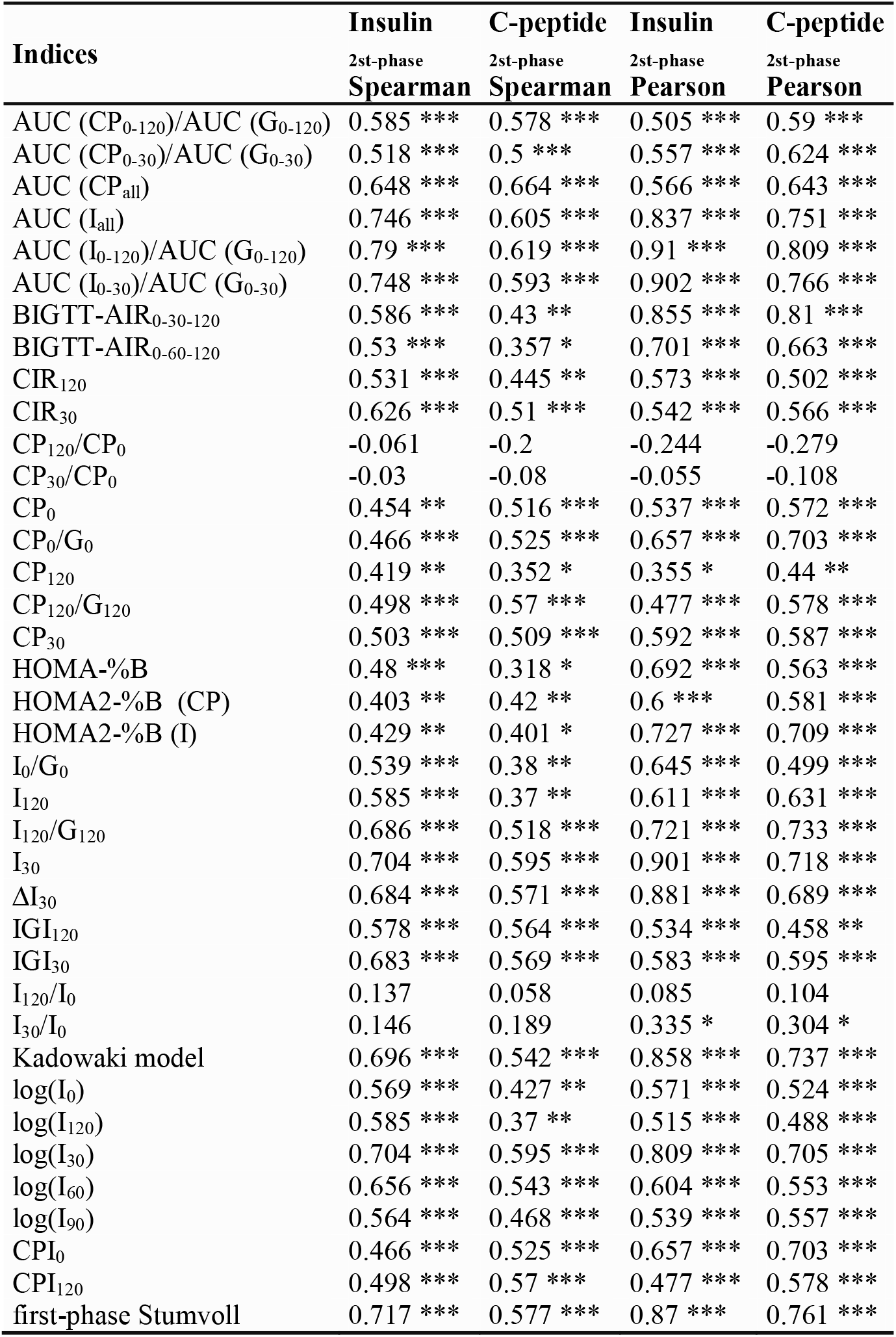
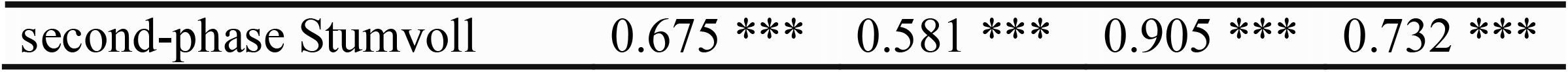
NGT-group(n=49). Correlation between fasting and OGTT-derived insulin secretion indices and second-phase insulin secretion measured with the hyperglycaemic clamp. (* p<0.05; **p<0.01; ***p<0.001)

**Figure 3.**
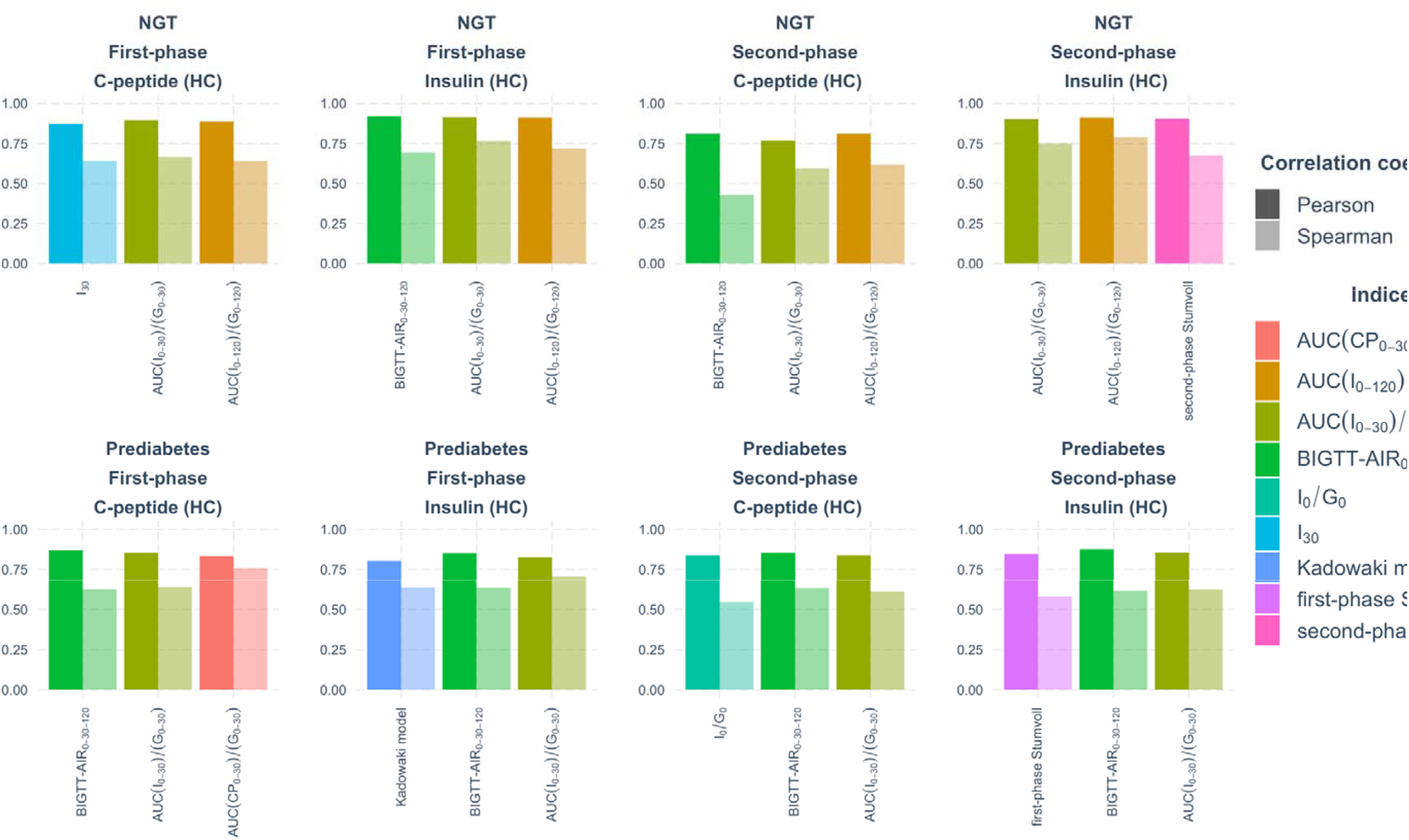
Top-indices by Spearman’s and Pearson’s correlation coefficients in NGT and prediabetes groups compared to first- and second phase hyperglycaemic clamp

According to both correlation coefficients, the strongest relationship between 1st-phase clamp- and OGTT-derived measurements in prediabetes correspond to AUC (I_0-30_)/AUC (G_0-30_), AUC (CP_0-30_)/AUC (G_0-30_), BIGTT-AIR_0-30-120_, Kadowaki model (Figure 3, Table 6.1).

**Table 6.1:** IFG and/or IGT -group (n=23). Correlation between fasting and OGTT-derived insulin secretion indices and first-phase insulin secretion measured with the hyperglycaemic clamp. (* p<0.05; **p<0.01; ***p<0.001)

| Indices | Insulin<sub>1st-phase</sub><br>Spearman | C-peptide<sub>1st-phase</sub><br>Spearman | Insulin<sub>1st-phase</sub><br>Pearson | C-peptide<sub>1st-phase</sub><br>Pearson |
| --- | --- | --- | --- | --- |
| AUC (CP<sub>0-120</sub>)/AUC (G<sub>0-120</sub>) | 0.605 ** | 0.659 *** | 0.589 ** | 0.655 *** |
| AUC (CP<sub>0-30</sub>)/AUC (G<sub>0-30</sub>) | 0.768 *** | 0.757 *** | 0.788 *** | 0.833 *** |
| AUC (CP<sub>all</sub>) | 0.657 ** | 0.646 ** | 0.477 * | 0.575 ** |
| AUC (I<sub>all</sub>) | 0.582 ** | 0.517 * | 0.51 * | 0.611 ** |
| AUC (I<sub>0-120</sub>)/AUC (G<sub>0-120</sub>) | 0.69 *** | 0.646 *** | 0.666 *** | 0.75 *** |
| AUC (I<sub>0-30</sub>)/AUC (G<sub>0-30</sub>) | 0.704 *** | 0.639 ** | 0.825 *** | 0.852 *** |
| BIGTT-AIR<sub>0-30-120</sub> | 0.634 ** | 0.626 ** | 0.851 *** | 0.868 *** |
| BIGTT-AIR<sub>0-60-120</sub> | 0.78 *** | 0.745 *** | 0.8 *** | 0.828 *** |
| CIR<sub>120</sub> | 0.601 ** | 0.633 ** | 0.33 | 0.42 |
| CIR<sub>30</sub> | 0.67 *** | 0.653 *** | 0.725 *** | 0.71 *** |
| CP<sub>120</sub>/CP<sub>0</sub> | -0.036 | 0.106 | -0.236 | -0.112 |
| CP<sub>30</sub>/CP<sub>0</sub> | 0.234 | 0.212 | 0.017 | 0.111 |
| CP<sub>0</sub> | 0.541 ** | 0.524 * | 0.731 *** | 0.749 *** |
| CP<sub>0</sub>/G<sub>0</sub> | 0.649 ** | 0.628 ** | 0.723 *** | 0.76 *** |
| CP<sub>120</sub> | 0.452 * | 0.57 ** | 0.229 | 0.339 |
| CP<sub>120</sub>/G<sub>120</sub> | 0.401 | 0.508 * | 0.129 | 0.2 |
| CP<sub>30</sub> | 0.764 *** | 0.718 *** | 0.722 *** | 0.785 *** |
| HOMA-%B | 0.688 *** | 0.608 ** | 0.733 *** | 0.784 *** |
| HOMA2-%B (CP) | 0.704 *** | 0.718 *** | 0.59 ** | 0.665 *** |
| HOMA2-%B (I) | 0.678 *** | 0.574 ** | 0.706 *** | 0.76 *** |
| I<sub>0</sub>/G<sub>0</sub> | 0.695 *** | 0.607 ** | 0.771 *** | 0.808 *** |
| I<sub>120</sub> | 0.633 ** | 0.636 ** | 0.399 | 0.529 ** |
| I<sub>120</sub>/G<sub>120</sub> | 0.623 ** | 0.615 ** | 0.437 * | 0.567 ** |
| I<sub>30</sub> | 0.669 *** | 0.6 ** | 0.775 *** | 0.813 *** |
| ΔI<sub>30</sub> | 0.63 ** | 0.593 ** | 0.753 *** | 0.79 *** |
| IGI<sub>120</sub> | 0.219 | 0.256 | 0.156 | 0.229 |
| IGI<sub>30</sub> | 0.625 ** | 0.615 ** | 0.461 * | 0.437 * |
| I<sub>120</sub>/I<sub>0</sub> | 0.146 | 0.205 | -0.08 | 0.031 |
| I<sub>30</sub>/I<sub>0</sub> | 0.268 | 0.268 | 0.064 | 0.089 |
| Kadowaki model | 0.636 ** | 0.608 ** | 0.803 *** | 0.817 *** |
| log(I<sub>0</sub>) | 0.618 ** | 0.511 * | 0.705 *** | 0.746 *** |
| log(I<sub>120</sub>) | 0.633 ** | 0.636 ** | 0.471 * | 0.586 ** |
| log(I<sub>30</sub>) | 0.669 *** | 0.6 ** | 0.651 *** | 0.705 *** |
| log(I<sub>60</sub>) | 0.561 ** | 0.503 * | 0.47 * | 0.509 * |
| log(I<sub>90</sub>) | 0.404 | 0.357 | 0.289 | 0.381 |
| CPI<sub>0</sub> | 0.649 ** | 0.628 ** | 0.723 *** | 0.76 *** |
| CPI<sub>120</sub> | 0.401 | 0.508 * | 0.129 | 0.2 |
| first-phase Stumvoll | 0.644 ** | 0.617 ** | 0.796 *** | 0.818 *** |
| second-phase Stumvoll | 0.615 ** | 0.559 ** | 0.772 *** | 0.809 *** |

The second-phase insulin and C-peptide strongly correlate with AUC (I_0-30_)/AUC (G_0-30_), AUC (I_0-120_)/AUC (G_0-120_), BIGTT-AIR_0-30-120_ in NGT (Figure 3, Table 5.2); and with I_0_/G_0_, AUC (I_0-30_)/AUC (G_0-30_), BIGTT-AIR_0-30-120_, first-phase Stumvoll in prediabetes (Figure 3, Table 6.2).

**Table 6.2:**
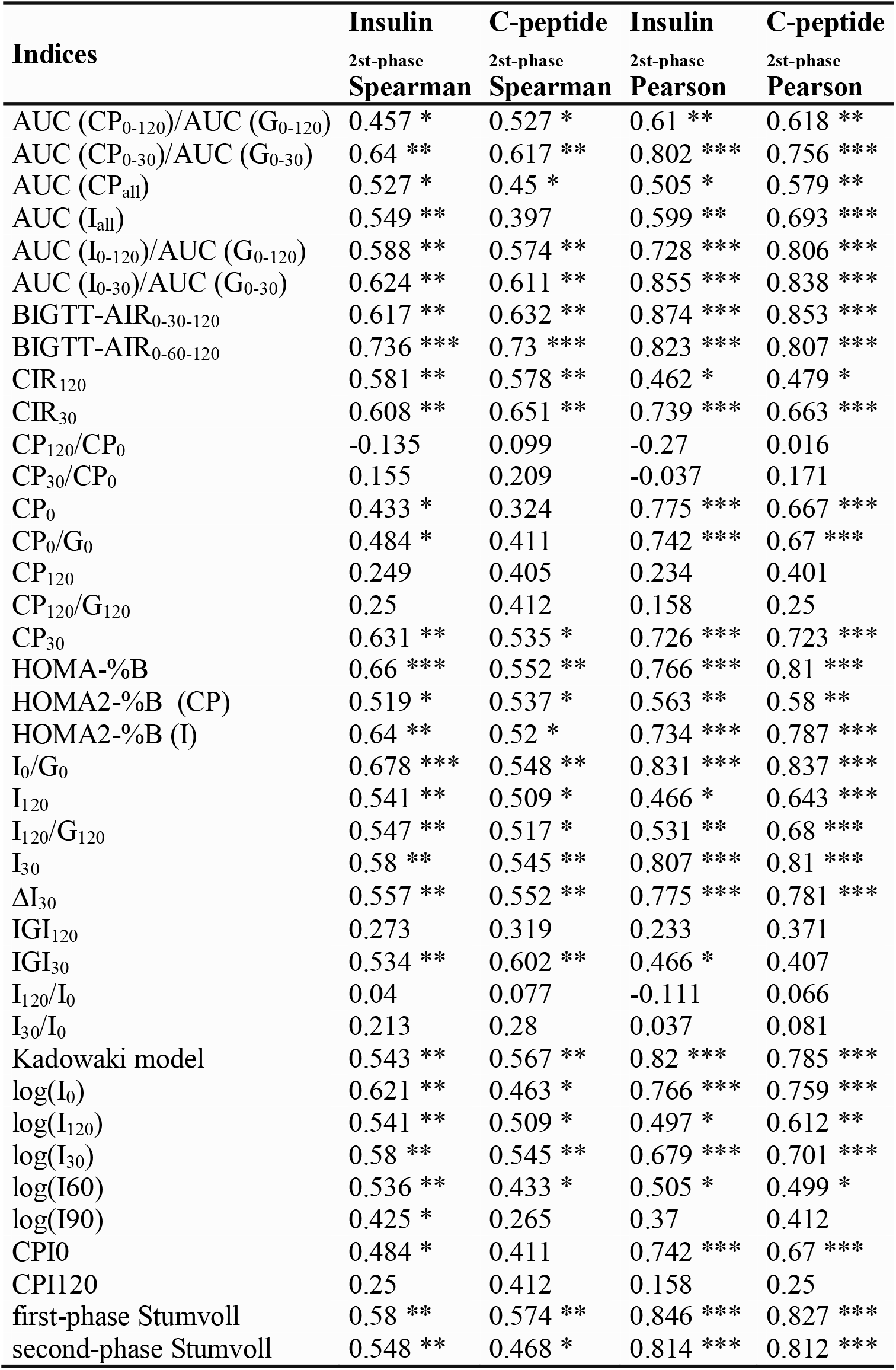
IFG and/or IGT -group (n=23). Correlation between fasting and OGTT-derived insulin secretion indices and second-phase insulin secretion measured with the hyperglycaemic clamp. (* p<0.05; **p<0.01; ***p<0.001)

| Indices | Insulin | C-peptide | Insulin | C-peptide |
| --- | --- | --- | --- | --- |
|  | 2st-phase<br>Spearman | 2st-phase<br>Spearman | 2st-phase<br>Pearson | 2st-phase<br>Pearson |
| AUC (CP<sub>0-120</sub>)/AUC (G<sub>0-120</sub>) | 0.457 * | 0.527 * | 0.61 ** | 0.618 ** |
| AUC (CP<sub>0-30</sub>)/AUC (G<sub>0-30</sub>) | 0.64 ** | 0.617 ** | 0.802 *** | 0.756 *** |
| AUC (CP<sub>all</sub>) | 0.527 * | 0.45 * | 0.505 * | 0.579 ** |
| AUC (I<sub>all</sub>) | 0.549 ** | 0.397 | 0.599 ** | 0.693 *** |
| AUC (I<sub>0-120</sub>)/AUC (G<sub>0-120</sub>) | 0.588 ** | 0.574 ** | 0.728 *** | 0.806 *** |
| AUC (I<sub>0-30</sub>)/AUC (G<sub>0-30</sub>) | 0.624 ** | 0.611 ** | 0.855 *** | 0.838 *** |
| BIGTT-AIR<sub>0-30-120</sub> | 0.617 ** | 0.632 ** | 0.874 *** | 0.853 *** |
| BIGTT-AIR<sub>0-60-120</sub> | 0.736 *** | 0.73 *** | 0.823 *** | 0.807 *** |
| CIR<sub>120</sub> | 0.581 ** | 0.578 ** | 0.462 * | 0.479 * |
| CIR<sub>30</sub> | 0.608 ** | 0.651 ** | 0.739 *** | 0.663 *** |
| CP<sub>120</sub>/CP<sub>0</sub> | -0.135 | 0.099 | -0.27 | 0.016 |
| CP<sub>30</sub>/CP<sub>0</sub> | 0.155 | 0.209 | -0.037 | 0.171 |
| CP<sub>0</sub> | 0.433 * | 0.324 | 0.775 *** | 0.667 *** |
| CP<sub>0</sub>/G<sub>0</sub> | 0.484 * | 0.411 | 0.742 *** | 0.67 *** |
| CP<sub>120</sub> | 0.249 | 0.405 | 0.234 | 0.401 |
| CP<sub>120</sub>/G<sub>120</sub> | 0.25 | 0.412 | 0.158 | 0.25 |
| CP<sub>30</sub> | 0.631 ** | 0.535 * | 0.726 *** | 0.723 *** |
| HOMA-%B | 0.66 *** | 0.552 ** | 0.766 *** | 0.81 *** |
| HOMA2-%B (CP) | 0.519 * | 0.537 * | 0.563 ** | 0.58 ** |
| HOMA2-%B (I) | 0.64 ** | 0.52 * | 0.734 *** | 0.787 *** |
| I<sub>0</sub>/G<sub>0</sub> | 0.678 *** | 0.548 ** | 0.831 *** | 0.837 *** |
| I<sub>120</sub> | 0.541 ** | 0.509 * | 0.466 * | 0.643 *** |
| I<sub>120</sub>/G<sub>120</sub> | 0.547 ** | 0.517 * | 0.531 ** | 0.68 *** |
| I<sub>30</sub> | 0.58 ** | 0.545 ** | 0.807 *** | 0.81 *** |
| ΔI<sub>30</sub> | 0.557 ** | 0.552 ** | 0.775 *** | 0.781 *** |
| IGI<sub>120</sub> | 0.273 | 0.319 | 0.233 | 0.371 |
| IGI<sub>30</sub> | 0.534 ** | 0.602 ** | 0.466 * | 0.407 |
| I<sub>120</sub>/I<sub>0</sub> | 0.04 | 0.077 | -0.111 | 0.066 |
| I<sub>30</sub>/I<sub>0</sub> | 0.213 | 0.28 | 0.037 | 0.081 |
| Kadowaki model | 0.543 ** | 0.567 ** | 0.82 *** | 0.785 *** |
| log(I<sub>0</sub>) | 0.621 ** | 0.463 * | 0.766 *** | 0.759 *** |
| log(I<sub>120</sub>) | 0.541 ** | 0.509 * | 0.497 * | 0.612 ** |
| log(I<sub>30</sub>) | 0.58 ** | 0.545 ** | 0.679 *** | 0.701 *** |
| log(I<sub>60</sub>) | 0.536 ** | 0.433 * | 0.505 * | 0.499 * |
| log(I<sub>90</sub>) | 0.425 * | 0.265 | 0.37 | 0.412 |
| CPI<sub>0</sub> | 0.484 * | 0.411 | 0.742 *** | 0.67 *** |
| CPI<sub>120</sub> | 0.25 | 0.412 | 0.158 | 0.25 |
| first-phase Stumvoll | 0.58 ** | 0.574 ** | 0.846 *** | 0.827 *** |
| second-phase Stumvoll | 0.548 ** | 0.468 * | 0.814 *** | 0.812 *** |

The highest correlation coefficients with first- and second-phase insulin secretion are shown by AUC (I_0-30_)/AUC (G_0-30_), BIGTT-AIR_0-30-120_, I_30_ and first-phase Stumvoll (Figure 4). The surrogate measures with the lowest correlation are CP_120_, log(I_120_) and log(I_90_).

**Figure 4.**
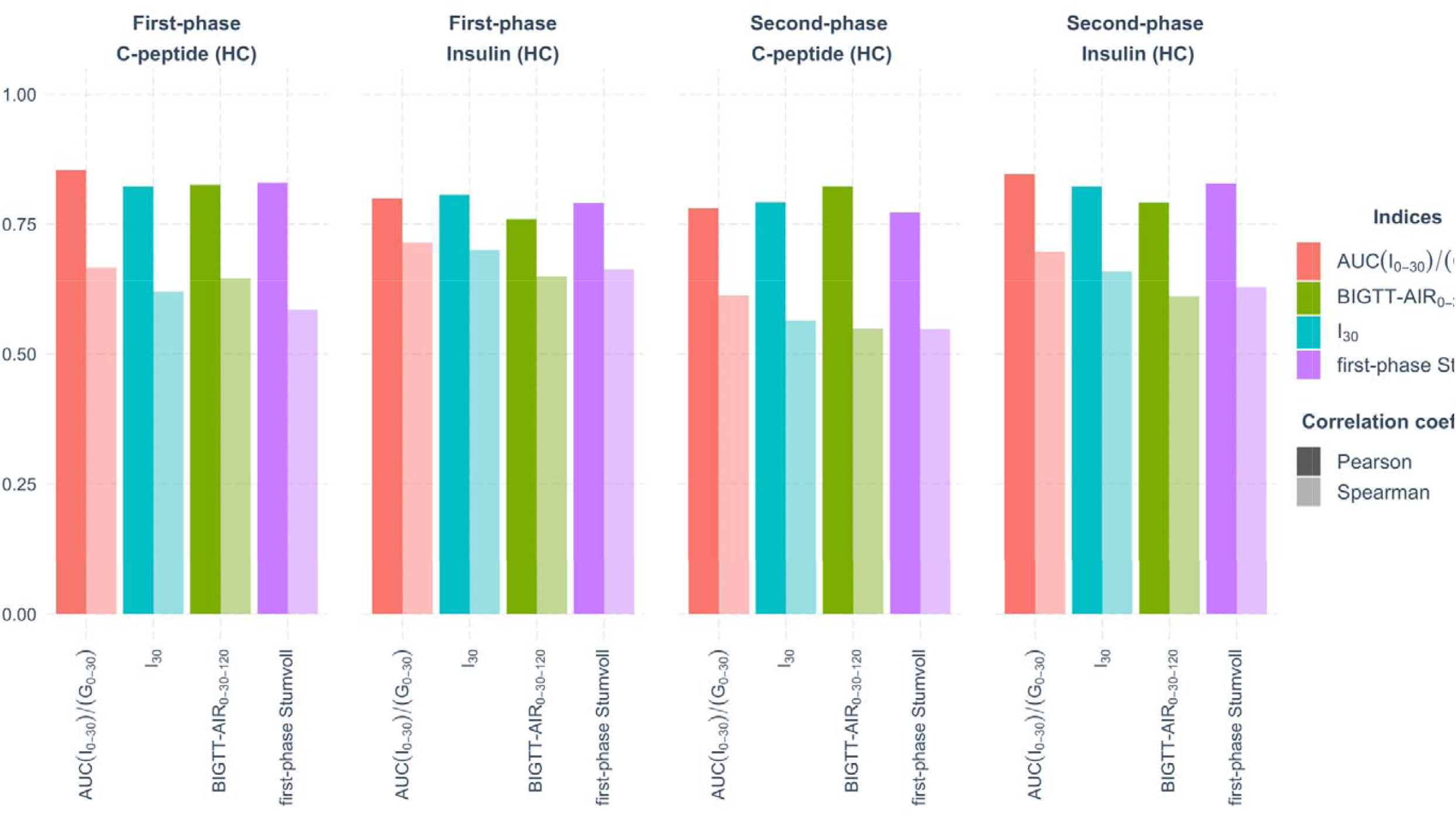
Top-indices by Spearman’s and Pearson’s correlation coefficients in the whole group (n=76) compared to first- and second phase hyperglycaemic clamp

Due to the small number of participants with test-diagnosed type 2 diabetes (n=4), they were excluded from this analysis.

Comparison of surrogate indices measuring insulin secretion using OGTT with the gold standard criteria revealed three indices (AUC (I_0-30_)/AUC (G_0-30_), first-phase Stumvoll, Kadowaki model) with the highest Pearson’s and Spearman’s coefficients for both groups (NGT and prediabetes) in two phases and during all tests (Figure 5). However, the CIR_120_ and IGI_120_ are the overall indices with the lowest significant correlation compared to the insulin and C-peptide gold standard measurements according to both correlations’ methods in the groups.

**Figure 5.**
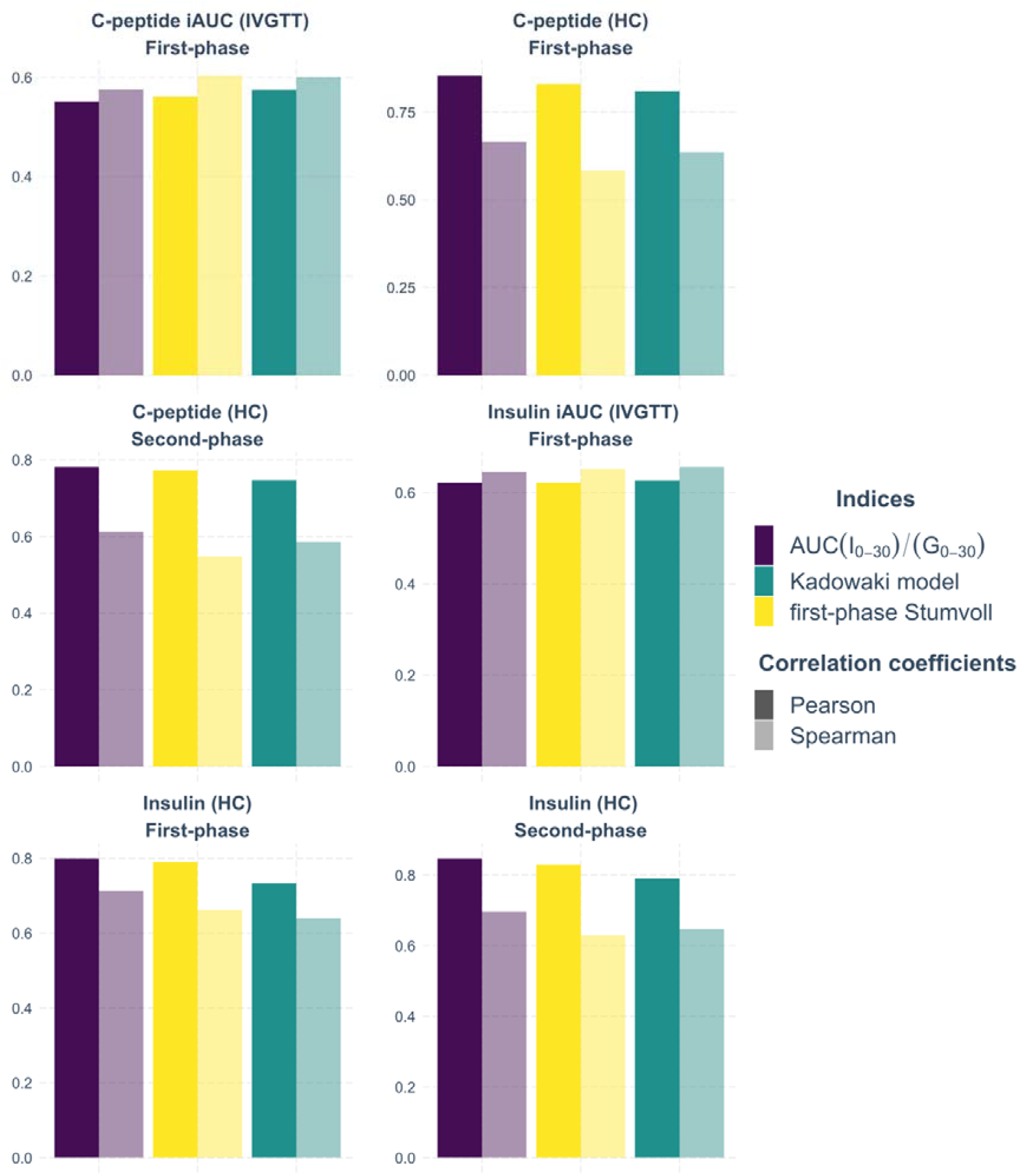
Indices with the highest Spearman’s and Pearson’s correlation coefficients in prediabetes and NGT groups in both study cohorts.

**Figure 6.**
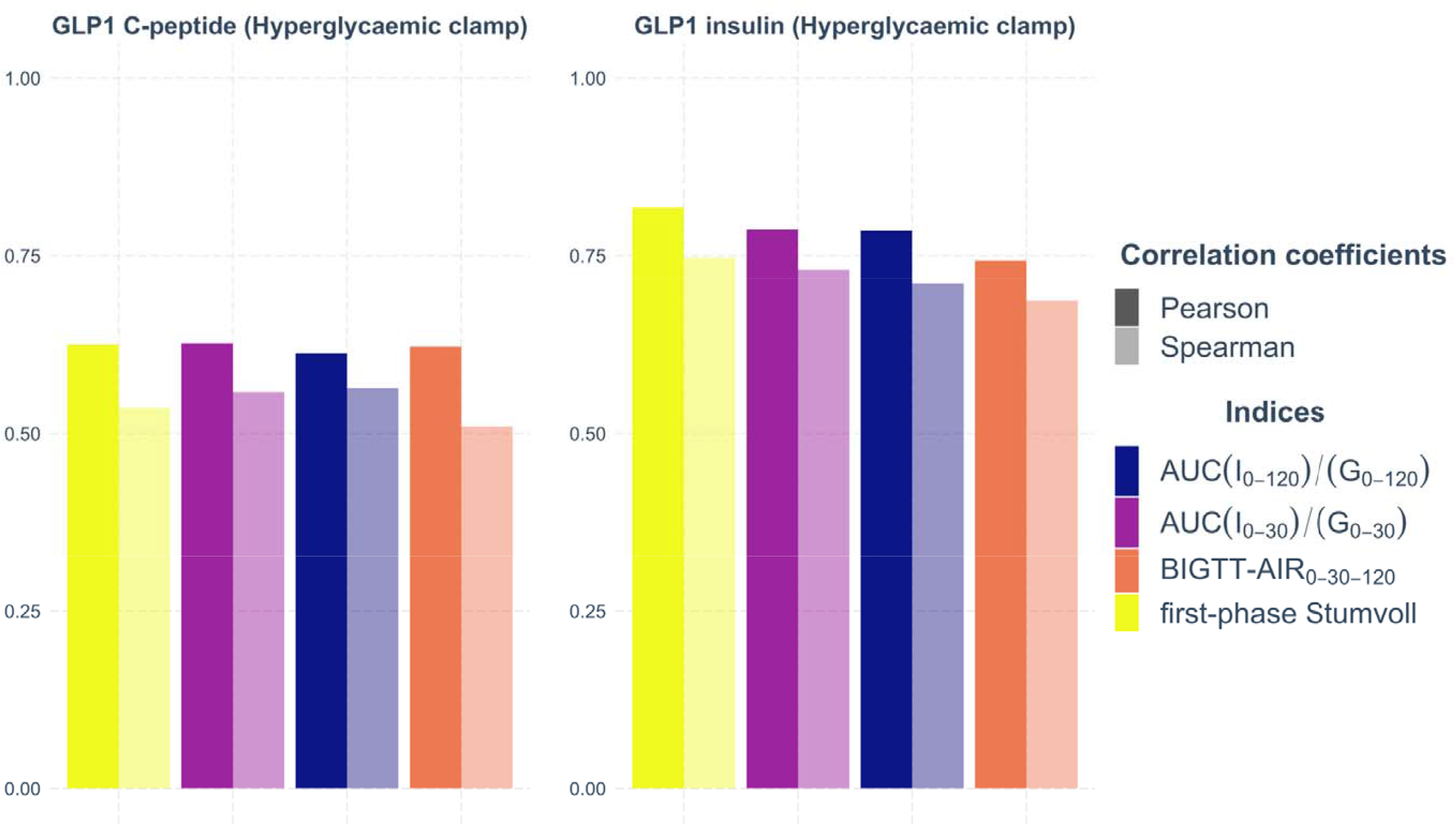
Top-indices by Spearman’s and Pearson’s correlation coefficients in a group of 76 participants compared to GLP-1 stimulated insulin secretion during hyperglycaemic clamp

### GLP-1-induced insulin (C-peptide) secretion

Using data from the modified hyperglycemic clamp, with a GLP-1 infusion phase^16^, we also tested the relationship between GLP-1 stimulated insulin release and OGTT-derived measurements. The vast majority of indices in NGT and prediabetes correlate significantly with clamp-derived GLP1-stimulated insulin secretion index (Table 7.a, Table 7.b). The strongest correlation belongs to first-phase Stumvoll, AUC (I_0-120_)/AUC (G_0-120_), AUC (I_0-30_)/AUC (G_0-30_), and BIGTT-AIR_0-60-120_ (Figure 7). The weakest correlation coefficients have CIR_120_ and CPI_120_.

**Table 7.a.** NGT-group(n=49). Correlation between fasting and OGTT-derived insulin secretion indices and GLP-1 insulin secretion measured with the hyperglycaemic clamp. (* p<0.05; **p<0.01; ***p<0.001)

| Indices | Insulin<sub>GLP1</sub><br>Spearman | C-peptide<sub>GLP1</sub><br>Spearman | Insulin<sub>GLP1</sub><br>Pearson | C-peptide<sub>GLP1</sub><br>Pearson |
| --- | --- | --- | --- | --- |
| AUC (CP<sub>0-120</sub>)/AUC (G<sub>0-120</sub>) | 0.525 *** | 0.38 ** | 0.511 *** | 0.45 ** |
| AUC (CP<sub>0-30</sub>)/AUC (G<sub>0-30</sub>) | 0.509 *** | 0.395 ** | 0.607 *** | 0.565 *** |
| AUC (CP<sub>all</sub>) | 0.484 *** | 0.343 * | 0.526 *** | 0.48 *** |
| AUC (I<sub>all</sub>) | 0.613 *** | 0.414 ** | 0.79 *** | 0.638 *** |
| AUC (I<sub>0-120</sub>)/AUC (G<sub>0-120</sub>) | 0.733 *** | 0.538 *** | 0.785 *** | 0.6 *** |
| AUC (I<sub>0-30</sub>)/AUC (G<sub>0-30</sub>) | 0.728 *** | 0.54 *** | 0.775 *** | 0.57 *** |
| BIGTT-AIR<sub>0-30-120</sub> | 0.631 *** | 0.466 *** | 0.715 *** | 0.6 *** |
| BIGTT-AIR<sub>0-60-120</sub> | 0.599 *** | 0.376 ** | 0.761 *** | 0.703 *** |
| CIR<sub>120</sub> | 0.435 ** | 0.32 * | 0.378 * | 0.266 |
| CIR<sub>30</sub> | 0.669 *** | 0.523 *** | 0.545 *** | 0.41 ** |
| CP<sub>120</sub>/CP<sub>0</sub> | -0.252 | -0.372 ** | -0.352 * | -0.364 * |
| CP<sub>30</sub>/CP<sub>0</sub> | -0.031 | -0.038 | -0.077 | -0.107 |
| CP<sub>0</sub> | 0.361 * | 0.304 * | 0.496 *** | 0.369 * |
| CP<sub>0</sub>/G<sub>0</sub> | 0.398 ** | 0.357 * | 0.678 *** | 0.691 *** |
| CP<sub>120</sub> | 0.182 | -0.023 | 0.32 * | 0.321 * |
| CP<sub>120</sub>/G<sub>120</sub> | 0.386 ** | 0.29 * | 0.409 ** | 0.415 ** |
| CP<sub>30</sub> | 0.445 ** | 0.355 * | 0.568 *** | 0.494 *** |
| HOMA-%B | 0.431 ** | 0.304 * | 0.674 *** | 0.59 *** |
| HOMA2-%B (CP) | 0.323 * | 0.291 * | 0.552 *** | 0.531 *** |
| HOMA2-%B (I) | 0.34 * | 0.344 * | 0.685 *** | 0.668 *** |
| I<sub>0</sub>/G<sub>0</sub> | 0.48 *** | 0.304 * | 0.623 *** | 0.407 ** |
| I<sub>120</sub> | 0.411 ** | 0.174 | 0.583 *** | 0.542 *** |
| I<sub>120</sub>/G<sub>120</sub> | 0.528 *** | 0.314 * | 0.64 *** | 0.568 *** |
| I<sub>30</sub> | 0.663 *** | 0.519 *** | 0.734 *** | 0.507 *** |
| ΔI<sub>30</sub> | 0.637 *** | 0.498 *** | 0.689 *** | 0.449 ** |
| IGI<sub>120</sub> | 0.568 *** | 0.541 *** | 0.295 | 0.247 |
| IGI<sub>30</sub> | 0.743 *** | 0.578 *** | 0.583 *** | 0.413 ** |
| I<sub>120</sub>/I<sub>0</sub> | -0.017 | -0.119 | -0.078 | -0.137 |
| I<sub>30</sub>/I<sub>0</sub> | 0.13 | 0.162 | 0.206 | 0.112 |
| Kadowaki model | 0.701 *** | 0.527 *** | 0.707 *** | 0.486 *** |
| log(I<sub>0</sub>) | 0.503 *** | 0.333 * | 0.644 *** | 0.556 *** |
| log(I<sub>120</sub>) | 0.411 ** | 0.174 | 0.461 *** | 0.319 * |
| log(I<sub>30</sub>) | 0.663 *** | 0.519 *** | 0.737 *** | 0.56 *** |
| log(I<sub>60</sub>) | 0.556 *** | 0.362 * | 0.627 *** | 0.489 *** |
| log(I<sub>90</sub>) | 0.319 * | 0.185 | 0.454 ** | 0.384 ** |
| CPI<sub>0</sub> | 0.398 ** | 0.357 * | 0.678 *** | 0.691 *** |
| CPI<sub>120</sub> | 0.386 ** | 0.29 * | 0.409 ** | 0.415 ** |
| first-phase Stumvoll | 0.739 *** | 0.573 *** | 0.815 *** | 0.617 *** |
| second-phase Stumvoll | 0.6 *** | 0.468 *** | 0.776 *** | 0.582 *** |

**Table 7.b.** IFG and/or IGT -group (n=23). Correlation between fasting and OGTT-derived insulin secretion indices and GLP-1 insulin secretion measured with the hyperglycaemic clamp. (* p<0.05; **p<0.01; ***p<0.001)

| **Indices** | **Insulin<sub>GLP1</sub>**<br>**Spearman** | **C-peptide<sub>GLP1</sub>**<br>**Spearman** | **Insulin<sub>GLP1</sub>**<br>**Pearson** | **C-peptide<sub>GLP1</sub>**<br>**Pearson** |
| --- | --- | --- | --- | --- |
| AUC (CP<sub>0-120</sub>)/AUC (G<sub>0-120</sub>) | 0.652 ** | 0.412 | 0.669 *** | 0.523 * |
| AUC (CP<sub>0-30</sub>)/AUC (G<sub>0-30</sub>) | 0.736 *** | 0.557 ** | 0.768 *** | 0.624 ** |
| AUC (CP<sub>all</sub>) | 0.49 * | 0.241 | 0.545 ** | 0.454 * |
| AUC (I<sub>all</sub>) | 0.483 * | 0.269 | 0.681 *** | 0.573 ** |
| AUC (I<sub>0-120</sub>)/AUC (G<sub>0-120</sub>) | 0.654 *** | 0.53 * | 0.772 *** | 0.698 *** |
| AUC (I<sub>0-30</sub>)/AUC (G<sub>0-30</sub>) | 0.725 *** | 0.517 * | 0.799 *** | 0.706 *** |
| BIGTT-AIR<sub>0-30-120</sub> | 0.66 *** | 0.49 * | 0.771 *** | 0.675 *** |
| BIGTT-AIR<sub>0-60-120</sub> | 0.682 *** | 0.519 * | 0.741 *** | 0.619 ** |
| CIR<sub>120</sub> | 0.608 ** | 0.38 | 0.529 * | 0.318 |
| CIR<sub>30</sub> | 0.777 *** | 0.642 ** | 0.702 *** | 0.61 ** |
| CP<sub>120</sub>/CP<sub>0</sub> | -0.119 | 0.033 | -0.169 | 0.047 |
| CP<sub>30</sub>/CP<sub>0</sub> | 0.238 | 0.317 | 0.048 | 0.332 |
| CP<sub>0</sub> | 0.489 * | 0.19 | 0.691 *** | 0.436 * |
| CP<sub>0</sub>/G<sub>0</sub> | 0.516 * | 0.269 | 0.677 *** | 0.464 * |
| CP<sub>120</sub> | 0.292 | 0.234 | 0.312 | 0.303 |
| CP<sub>120</sub>/G<sub>120</sub> | 0.5 * | 0.321 | 0.252 | 0.185 |
| CP<sub>30</sub> | 0.606 ** | 0.456 * | 0.685 *** | 0.602 ** |
| HOMA-%B | 0.565 ** | 0.349 | 0.755 *** | 0.649 *** |
| HOMA2-%B (CP) | 0.582 ** | 0.396 | 0.553 ** | 0.457 * |
| HOMA2-%B (I) | 0.543 * | 0.296 | 0.729 *** | 0.634 ** |
| I<sub>0</sub>/G<sub>0</sub> | 0.582 ** | 0.329 | 0.805 *** | 0.66 *** |
| I<sub>120</sub> | 0.339 | 0.358 | 0.584 ** | 0.561 ** |
| I<sub>120</sub>/G<sub>120</sub> | 0.529 * | 0.416 * | 0.656 *** | 0.587 ** |
| I<sub>30</sub> | 0.642 ** | 0.488 * | 0.759 *** | 0.689 *** |
| ΔI<sub>30</sub> | 0.609 ** | 0.487 * | 0.726 *** | 0.676 *** |
| IGI<sub>120</sub> | 0.358 | 0.155 | 0.374 | 0.316 |
| IGI<sub>30</sub> | 0.718 *** | 0.581 ** | 0.484 * | 0.473 * |
| I<sub>120</sub>/I<sub>0</sub> | -0.113 | 0.089 | -0.073 | 0.04 |
| I<sub>30</sub>/I<sub>0</sub> | 0.341 | 0.343 | 0.064 | 0.199 |
| Kadowaki model | 0.686 *** | 0.533 ** | 0.755 *** | 0.681 *** |
| log(I<sub>0</sub>) | 0.544 ** | 0.262 | 0.75 *** | 0.593 ** |
| log(I<sub>120</sub>) | 0.339 | 0.358 | 0.509 * | 0.507 * |
| log(I<sub>30</sub>) | 0.642 ** | 0.488 * | 0.684 *** | 0.634 ** |
| log(I<sub>60</sub>) | 0.411 | 0.204 | 0.481 * | 0.359 |
| log(I<sub>90</sub>) | 0.339 | 0.11 | 0.461 * | 0.335 |
| CPI<sub>0</sub> | 0.516 * | 0.269 | 0.677 *** | 0.464 * |
| CPI<sub>120</sub> | 0.5 * | 0.321 | 0.252 | 0.185 |
| first-phase Stumvoll | 0.757 *** | 0.545 ** | 0.831 *** | 0.706 *** |
| second-phase Stumvoll | 0.544 ** | 0.345 | 0.758 *** | 0.665 *** |

## DISCUSSION

In this study, we compared fasting and/or OGTT-based insulin secretion markers to gold-standard measurements. Our study revealed that most surrogate indices of insulin secretion correlated significantly with dynamic measurements assessed from IVGTT and hyperglycaemic clamp. The substantial difference between Pearson’s and Spearman’s correlation coefficients for some surrogate indices can be attributed to variables with skewed distributions.

The overall highest correlation coefficients calculated by both Pearson’s and Spearman’s tests were found for AUC (I_0-30_)/AUC (G_0-30_), first-phase Stumvoll and Kadowaki model. These indices have high correlation coefficients with gold standard measures, obtained both from insulin and C-peptide measurements, both with IVGTT and hyperglycaemic clamp. All three indices reflect insulin release and comprise OGTT-derived insulin and glucose levels at minutes 0 and 30 and predict insulin secretion abnormalities better than 120-minute-based indices (e.g. CP_120_, CIR_120_ and IGI_120_). The somewhat weaker correlation with 120-minute indices can probably be due to changes in the incretin axis caused by oral intake of glucose^27^. The high correlation of AUC (I_0-120_)/AUC (G_0-120_) and BIGTT-AIR_0-60-120_ with stimulated GLP-1 insulin secretion in our analysis argues for a longer assessment during OGTT when addressing incretin effects. The strong relationship between incretin-stimulated response and AUC (I_0-30_)/AUC (G_0-30_), first-phase Stumvoll in all experiments stresses the robustness of these surrogate indices to estimate insulin secretion. Another possible contributor to a comparably better correlation of short-term insulin/C-peptide assessment during OGTT with gold-standard measures could be a diverging influence of insulin clearance over time^9^.

Parenteral and oral glucose loading leads to stimulation of glucose with the involvement of various physiological processes. Oral glucose ingestion activates complex mechanisms, including an incretin-related cascade and even the brain^28^ that all in concert control insulin release Taken together, we here determined which indices can most reliably estimate insulin secretion from fasting measures or OGTT. Given the importance of assessing insulin secretion for the classification of patients in novel subgroups of prediabetes^5^ and diabetes^4^, also for potential therapeutic considerations^29^, selection of appropriate estimates from OGTTs is crucial in research and could also play a role in future clinical practice.

## Data Availability

The datasets analyzed during the current study are not publicly available owing to ethical regulations, but are available from the corresponding author on reasonable request.

## ACKNOWLEDGMENTS

We thank all the volunteers of the study for their participation. We acknowledge support from the Helmholtz Center Munich, Deutsche Forschungsgemeinschaft, the German Center for Diabetes Research (DZD e.V.), and the Open Access Publishing Foundation of the University of Tübingen.

## AUTHOR CONTRIBUTIONS

KP, RW and MH designed the analysis strategy, supervised the project and wrote the manuscript. RR, YC, JO, AB, AP, and AF contributed to the interpretation of the data and edited the manuscript.

## FUNDING

This work was supported in part by grant 01GI0925 from the German Federal Ministry of Education and Research (BMBF) to the German Center for Diabetes Research (DZD).

## CONFLICT OF INTEREST STATEMENT

Outside of the current work, R.W. does report lecture fees from Novo Nordisk and Sanofi. He served on the advisory board of Akcea Therapeutics, Daiichi Sankyo, Sanofi and NovoNordisk. Outside of the current work, A.F. reports lecture fees and advisory board membership from Sanofi, Novo Nordisk, Eli Lilly, and AstraZeneca. Outside of the current work, M.H. reports research grants from Boehringer Ingelheim and Sanofi (both to the University Hospital of Tübingen), advisory board for Boehringer Ingelheim, and lecture fees from Boehringer Ingelheim, Sanofi, Novo Nordisk, Eli Lilly and Merck Sharp & Dohme.

None of the other authors report a conflict of interest.

## Notes

### Author Declarations

The study protocols were approved by the Ethical Committee of the Medical Faculty of the University of Tuebingen.

